# High HIV prevalence, declining mortality: contrasting epidemic trajectories in the Amazon and the Guianas

**DOI:** 10.64898/2026.11.26.26369050

**Authors:** Mathieu Nacher, Aude Lucarelli, Abire Bawa, Pierre Couppié, Antoine Adenis

## Abstract

**Background:** The Guiana Shield has the highest HIV prevalence in South America. Advanced HIV remains a public health problem and the diagnosis and treatment of disseminated histoplasmosis is still lagging. French Guiana, a French territory where antiretroviral treatment is free and universal, has long had the most severe epidemic of any French region, but it is not a location in the Global Burden of Disease (GBD) study.

**Methods:** We extracted GBD 2023 age-standardized HIV/AIDS death and DALY rates (both sexes, 1990–2023) for France, the Guiana Shield, Andean–Amazon countries including Peru, and the nine states of the Brazilian Legal Amazon. For French Guiana and mainland France we took 2001–2024 underlying-cause deaths (category 1.2, ICD-10 B20–B24) from CépiDc-Inserm by sex and 10-year age group, with INSEE populations, and computed directly standardized rates and age-adjusted Poisson trends. French Guiana was placed on the GBD scale by a 20,000-draw Monte Carlo model calibrated on GBD France ÷ CépiDc mainland France.

**Results:** In 2019–2023, GBD mortality was highest in Guyana (19.0 per 100,000) and Suriname (16.3), against 3.9–8.6 in the Brazilian Amazon states. French Guiana recorded 453 AIDS deaths in 2001–2024. Its world-standardized rate fell from 22.7 per 100,000 in 2001–2005 to 4.0 (95% CI 3.0–5.2) in 2019–2023, −9.7% a year (95% CI −11.1 to −8.4). Mainland France fell in parallel, so the gap did not close: French Guiana was 16 times the mainland rate in 2001–2005 and 19 times in 2019–2023. The excess was larger in women (×23) than in men (×17). On the GBD scale (calibration factor k = 2.12), French Guiana’s 2019–2023 rate was 8.3 (6.0–11.2) per 100,000, 19 times France, level with Amazonas and Pará, above the Andean countries and half that of Suriname and Guyana. DALYs were 499 (377–652) per 100,000. By sex, the GBD-equivalent rate was 10.7 per 100,000 in men (17× France) and 6.3 in women (23× France). Across the region, the male:female ratio of HIV prevalence ranged from 0.87 in Suriname to 3.5 in Peru; French Guiana’s male:female mortality ratio (1.7) was close to those of Guyana and Suriname and below France’s (2.3). Assuming that 20–40% of HIV deaths are due to histoplasmosis, about 7 (4–10) HIV-associated histoplasmosis deaths a year occur in French Guiana, 2.5 per 100,000, against 4.9 in Suriname and 5.7 in Guyana.

**Conclusions:** Universal antiretroviral treatment cut AIDS mortality in French Guiana by more than 80%, faster than in its neighbors. Its sex pattern is that of the gender-balanced epidemics of the Guiana Shield, not that of mainland France, which explains why women carry the largest relative excess. Finding undiagnosed infections earlier and retaining them in care to achieve the 95 95 95 target remains the priority.

## Introduction

Adult HIV prevalence in French Guiana, Guyana and Suriname, 1.0–1.5%, is among the highest on the American continent, and the epidemic there is driven by Caribbean as well as pandemic subtype B lineages that circulate across the borders.[1,2] The phylogenetic studies show that the historical viral circulation in the region is complex bridging the Caribbean epidemic mostly due to B_Car_ viruses with the South American mainly due to B _Pandemic_ viruses. Hence it has been shown that while B _Pandemic_ is predominant in Brazil the states where the B _Car_ virus proportion is highest are the Amazonian states where Garimpeiros come from suggesting a bridging role between the Caribbean and Amazonian epidemic.[3] In French Guiana, HIV prevalence among pregnant women has exceeded 1% for more than two decades, and AIDS incidence was ten times the national average.[4] Transmission is mainly heterosexual, men and women are equally affected, and sex work, crack cocaine use and multiple concurrent partnerships are major drivers.[5]

French Guiana has the medical resources of France: all antiretrovirals, free care regardless of residence status, and a cohort that has documented the epidemic since 1992. By 2018, 91% of patients in care were on treatment and 94% of them had an undetectable viral load and in 2025 the 95*95*95 objectives were attained.[6,7] Yet the share of patients diagnosed at an advanced stage has long resisted progress[8], and disseminated histoplasmosis, often mistaken for tuberculosis, remains the leading AIDS-defining infection and cause of AIDS death.[9,10] But French Guiana should not be isolated from its Amazonian situation. Populations are highly mobile through the region and the Amazonian pathogen ecology influences the presentation of opportunistic infections in persons with advanced HIV.[11,12]

Amazonia and the Guiana Shield form a single ecological and epidemiological entity marked by isolation, difficult transport, remote and often poor populations, and illegal gold mining, sex work and trafficking that ignore borders. Yet it is divided among nine countries and territories whose health and research systems rarely meet. Studying it as a single entity may reveal common problems and gaps that are difficult to apprehend from national perspectives alone. Because GBD pools French Guiana with France, its HIV burden is invisible in the comparative tools used for the Amazon region. Moreover, UNAIDS estimates on HIV programmatic aspects and milestones still project concerning numbers.[13] French Guiana occupies a distinctive position in this regional comparison. Geographically and epidemiologically, it belongs to the Amazon and the Guiana Shield, sharing cross-border population movements, social vulnerabilities and infectious disease exposures with its neighbors. Administratively, however, it is part of France and benefits from the same national healthcare and social protection framework, making mainland France a relevant benchmark for assessing persistent inequalities within a single country. Comparing French Guiana with both mainland France and its Amazonian neighbors therefore provides complementary perspectives: the former reveals inequalities despite a shared national health system, while the latter helps explore how HIV outcomes differ across geographically connected populations served by health systems with markedly different resources and organization.

Our aim was thus to describe HIV/AIDS mortality across the Amazon and the Guianas from GBD 2023, to integrate French Guiana in this framework and make regional comparisons..

## Methods

### Design and data

This is an ecological descriptive study. We downloaded GBD 2023 age-standardized HIV/AIDS death, YLL, YLD and DALY rates per 100,000 (both sexes, 1990–2023, with 95% uncertainty intervals), for both sexes and for men and women separately, together with HIV prevalence and incidence rates by sex, for France (reference), Suriname, Guyana, Venezuela, Colombia, Ecuador, Bolivia, Peru and the Brazilian states of Acre, Amapá, Amazonas, Pará, Rondônia, Roraima, Tocantins Maranhão, and Mato Grosso.[14]

For French Guiana and mainland France we took deaths with HIV disease as the underlying cause (European Short List category 1.2, ICD-10 B20–B24) from the CépiDc-Inserm open-data tool, 2001–2024, by sex and 10-year age group.[15] French Guiana counts came directly. For mainland France the tool returned crude rates by age; we rebuilt counts as rate × INSEE person-years. The rebuilt totals matched the published all-age counts within 0.3% on average (range of annual ratios 0.90–1.06). Tuberculosis, viral hepatitis and other infectious diseases were extracted as a comparison panel.

Person-years came from INSEE 1 January estimates by region, sex and 5-year age group (mean of 1 January of y and y+1); 2024, which has no INSEE estimate yet, was back-calculated from CépiDc crude rates for French Guiana and not available for mainland France.

### Analysis

We computed directly standardized rates over nine age groups with the WHO world standard (main), the European 2013 and Segi standards, with Fay–Feuer 95% CIs [8], for 5-year periods, for both sexes and for men and women separately. We validated the reconstruction against the rates CépiDc publishes. Trends were estimated with an age-adjusted Poisson model with log person-years as offset.

### GBD-equivalent estimate for French Guiana

We used 2019–2023 age-specific deaths and person-years for French Guiana. Each of 20,000 Monte Carlo draws drew age-specific rates from a Gamma(deaths) distribution; age groups with zero deaths stayed at zero. Each draw also multiplied the WHO world-standardized rate by a calibration factor k, which brings vital-registration rates onto the GBD scale. k = GBD France HIV/AIDS death rate (drawn from its 95% UI) ÷ the CépiDc rate for mainland France computed the same way (both sexes, 2019–2023, INSEE person-years). k captures what GBD adds to certified underlying causes, notably HIV deaths certified as an opportunistic infection or as an ill-defined cause. Each draw computed YLLs from deaths by age and a GBD-type reference life table, rescaled so that the YLL per death matches GBD France. Finally each draw set YLDs to GBD France YLDs times a triangular multiplier, Triangular(2.5, 4, 6), reflecting adult HIV prevalence of 1.1–1.5% in French Guiana against about 0.3% in mainland France.[6]

We report medians with 2.5th–97.5th percentile intervals, and the share of draws in which French Guiana exceeded each GBD location. The model was run for both sexes and then separately for men and women, each with its own k (sex-specific GBD France ÷ sex-specific CépiDc mainland rate) and sex-specific INSEE person-years.

### Simulation of histoplasmosis-attributable deaths

Histoplasmosis is not a separate cause in GBD, and HIV deaths due to histoplasmosis are coded within B20–B24 (B20.5) in vital registration, so neither source isolates them. We therefore simulated histoplasmosis-attributable HIV deaths as the HIV/AIDS death rate multiplied by an attributable fraction. The fraction was drawn from Triangular(20%, 30%, 40%), bracketing published evidence from the region: histoplasmosis up to one-fifth of AIDS deaths in Latin America[16], a major contributor to 22% of deaths in HIV-positive patients autopsied in Manaus[17], and proven or probable histoplasmosis in 40% of hospitalized people with HIV who died within 30 days in Suriname and French Guiana.[18] For GBD locations, the death rate was drawn from a log-normal distribution matching its 2019–2023 95% UI; for French Guiana we used the 20,000 GBD-equivalent draws. YLLs were computed the same way, assuming the same age distribution as all HIV deaths. In sensitivity analyses the fraction was Uniform(10%, 20%) and Uniform(30%, 45%). To express the burden in deaths a year, we multiplied each location’s age-standardized rate by its approximate 2022–2023 population (IBGE 2022 census for the Brazilian states; national estimates elsewhere); totals were summed draw by draw, with the attributable fraction shared across other locations within each draw. Because histoplasmosis diagnosis and treatment have evolved differently across territories, we complemented the regional attributable-fraction model with scenarios informed by published local case-fatality data. For French Guiana, we used the reported mean annual number of diagnosed histoplasmosis episodes during 2012–2019 (approximately 16) and the corresponding early case-fatality proportion of 3.9%.[12] For Suriname, we considered a historical 30-day case-fatality proportion of 17.5%, reported in a prospective hospital-based study conducted in French Guiana and Suriname during 2013–2015. [18] As this estimate was derived from the combined cohort, it was treated as a historical proxy rather than a Suriname-specific estimate. Expected early deaths were calculated as the annual number of histoplasmosis episodes multiplied by the corresponding case-fatality proportion. Where reliable population-based episode counts were unavailable, calculations were presented as conditional scenarios rather than national mortality estimates. These analyses were distinguished from the original model estimating the proportion of all HIV/AIDS deaths attributable to histoplasmosis.

### Sex differences across territories

For each GBD location we computed male:female ratios of the 2019–2023 age-standardized incidence, prevalence and death rates, and, as a rough indicator of lethality, deaths per 1,000 people living with HIV (ratio of the age-standardized death and prevalence rates). We followed the male:female prevalence ratio from 1990 to 2023.

### Prevalence and incidence

We mapped 2019–2023 mean age-standardized HIV prevalence and incidence rates. Both-sexes rates were approximated by the mean of the male and female rates. For French Guiana, which GBD does not cover, we took the number of people living with HIV (diagnosed plus undiagnosed) estimated by the ECDC HIV Modelling Tool for 2019–2023 [ref: ECDC HIV Modelling Tool] and divided it by the INSEE population on 1 January 2023 (301,099), giving a crude prevalence. New infections were approximated as the mean annual change in people living with HIV in 2020–2023 plus an assumed 30–60 deaths a year among them (10–22 HIV deaths on the CépiDc and GBD-equivalent scales, plus deaths from other causes); this is an indicative figure, not a modelled incidence.

## Results

### Regional picture from GBD 2023

In 2019–2023, GBD age-standardized HIV/AIDS mortality was highest in Guyana, 19.0 per 100,000 (95% UI 16.5–21.5), and Suriname, 16.3 (13.7–18.8) (Table 1). The Brazilian Amazon states ranged from 3.9 in Acre to 8.6 in Pará, with Amazonas, Pará, Maranhão (7.5), Roraima and Amapá at 7–9 and Mato Grosso at 6.1. Ecuador, Colombia, Bolivia, Peru (5.4) and Venezuela were at 4.7–5.7. France was at 0.45. YLLs made up 87–94% of DALYs in the Amazonian locations.

**Table 1.** HIV/AIDS mortality per 100,000 (age-standardized, GBD 2023, both sexes): 2019–2023 means, peak and change.

| Location | Deaths per 100,000, 2019–2023 (95% UI) | DALYs per 100,000 | YLL share (%) | Deaths 1990 | Peak (year) | Deaths 2023 | Change peak→2023 (%) |
|---|---|---|---|---|---|---|---|
| Guyana | 19.0 (16.5–21.5) | 1,013 | 92 | 14.7 | 53.7 (2001) | 19.5 | -64 |
| Suriname | 16.3 (13.7–18.8) | 868 | 93 | 11.1 | 30.7 (2003) | 16.2 | -47 |
| Pará (BR) | 8.6 (8.0–9.3) | 469 | 89 | 2.9 | 9.2 (2015) | 8.3 | -10 |
| Amazonas (BR) | 8.6 (7.9–9.4) | 471 | 88 | 3.0 | 9.4 (2016) | 8.4 | -10 |
| Maranhão (BR) | 7.5 (6.9–8.0) | 404 | 89 | 1.7 | 8.8 (2015) | 7.1 | -20 |
| Roraima (BR) | 7.4 (6.6–8.3) | 389 | 92 | 4.8 | 8.0 (2003) | 7.2 | -10 |
| Amapá (BR) | 7.1 (6.2–7.9) | 367 | 94 | 3.0 | 7.2 (2022) | 6.8 | -6 |
| Mato Grosso (BR) | 6.1 (5.6–6.6) | 313 | 91 | 3.4 | 7.5 (2008) | 5.9 | -21 |
| Venezuela | 5.7 (4.8–6.8) | 320 | 91 | 3.2 | 8.1 (2016) | 5.0 | -39 |
| Rondônia (BR) | 5.7 (5.1–6.3) | 286 | 92 | 3.0 | 5.9 (2022) | 5.7 | -3 |
| Peru | 5.4 (4.8–6.2) | 289 | 94 | 1.3 | 6.9 (2002) | 4.4 | -36 |
| Bolivia | 5.4 (3.0–8.4) | 268 | 94 | 0.2 | 24.6 (2007) | 4.6 | -81 |
| Colombia | 5.0 (4.6–5.3) | 267 | 91 | 2.2 | 6.5 (2006) | 4.6 | -30 |
| Ecuador | 4.7 (4.3–5.1) | 264 | 90 | 1.1 | 7.5 (2006) | 4.4 | -41 |
| Tocantins (BR) | 4.7 (4.2–5.2) | 241 | 93 | 1.5 | 4.9 (2022) | 4.6 | -5 |
| Acre (BR) | 3.9 (3.4–4.4) | 202 | 93 | 3.5 | 5.5 (1996) | 3.6 | -34 |
| French Guiana (GBD-equivalent) | 8.3 (6.0–11.2) | 499 | 81 | – | – | – | – |
| France (reference) | 0.45 (0.38–0.52) | 43 | 45 | 4.6 | 7.8 (1994) | 0.45 | -94 |

*French Guiana: GBD-equivalent estimate (median and 95% UI of 20,000 draws), CépiDc 2019–2023 deaths scaled by k = GBD France ÷ CépiDc mainland France (median 2.12). The observed CépiDc rate was 4.0 (3.0–5.2)*

The trajectories differed sharply (Figure 1). Guyana peaked at 54 per 100,000 in 2001 and fell by 64% to 2023; Suriname peaked at 31 in 2003 and fell by 47%. Bolivia’s estimates rose from almost nothing to 25 in 2007 and fell by 81%, with very wide UIs. In the Brazilian Amazon, by contrast, mortality rose slowly throughout and in 2023 was still within 3–10% of its peak in Pará, Amazonas, Roraima, Amapá, Tocantins and Rondônia. Maranhão rose from 1.7 in 1990 to 8.8 in 2015 and was 20% lower in 2023; Mato Grosso, already at 3.4 in 1990, rose to about 7 by the mid-1990s and stayed there until 2008 and was 21% lower in 2023. Peru peaked at 6.9 in 2002, fell to about 4 in the early 2010s, rose again to 6.2 in 2019 and stood at 4.4 in 2023.

**Figure 1.**
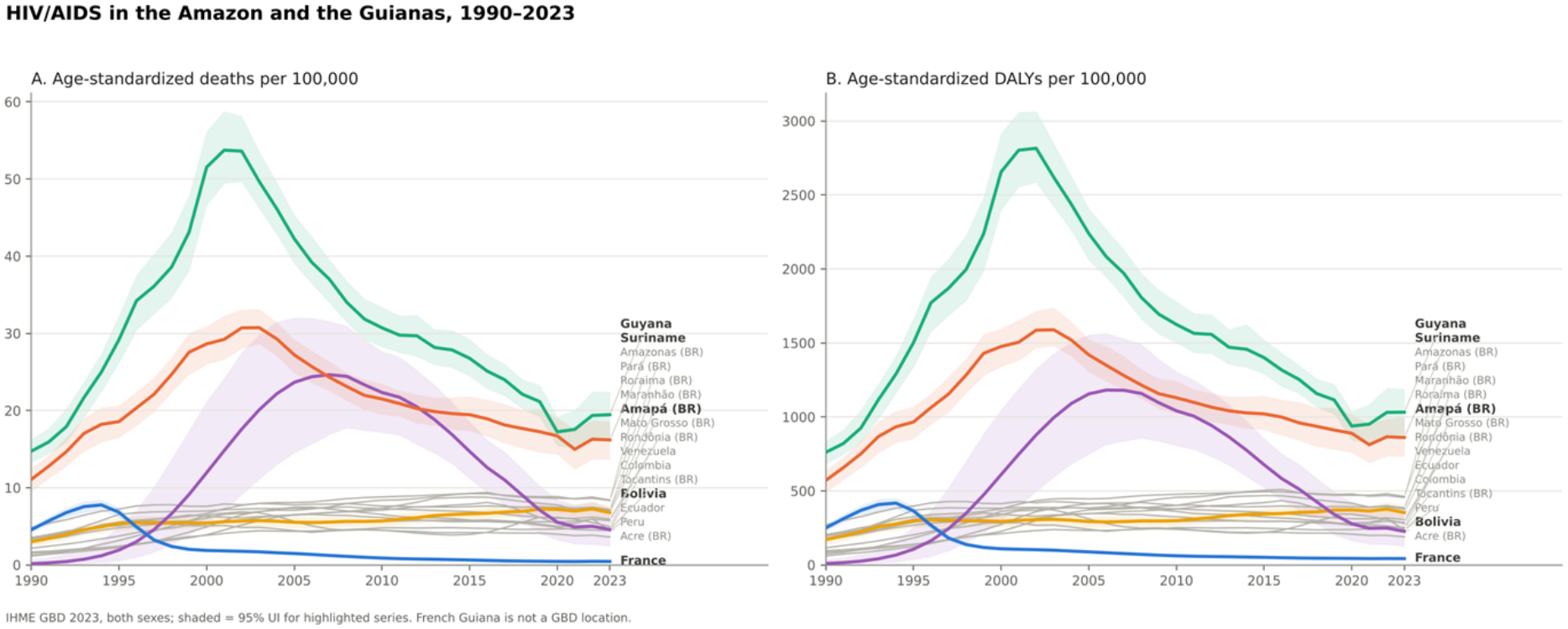
Age-standardized HIV/AIDS deaths (A) and DALYs (B) per 100,000, 1990–2023 (GBD 2023, both sexes). Shaded bands: 95% UIs for highlighted locations. French Guiana is not a GBD location.

### French Guiana and mainland France (CépiDc)

Our reconstruction matched CépiDc’s published sex-specific European-standard rates for French Guiana (correlation ≥ 0.998; mean absolute difference 0.44 per 100,000).

French Guiana recorded 453 AIDS deaths in 2001–2024: 34–38 a year in 2001–2005, and 6–13 a year since 2015. The world-standardized rate fell from 22.7 per 100,000 (95% CI 19.3–26.6) in 2001–2005 to 4.0 (3.0–5.2) in 2019–2023, a fall of 82%. The age-adjusted decline was −9.7% a year (95% CI −11.1 to −8.4). The steepest drop came in 2006, and the rate has been roughly flat at 2.4–5.3 since 2016 (Figure 2A).

**Figure 2.**
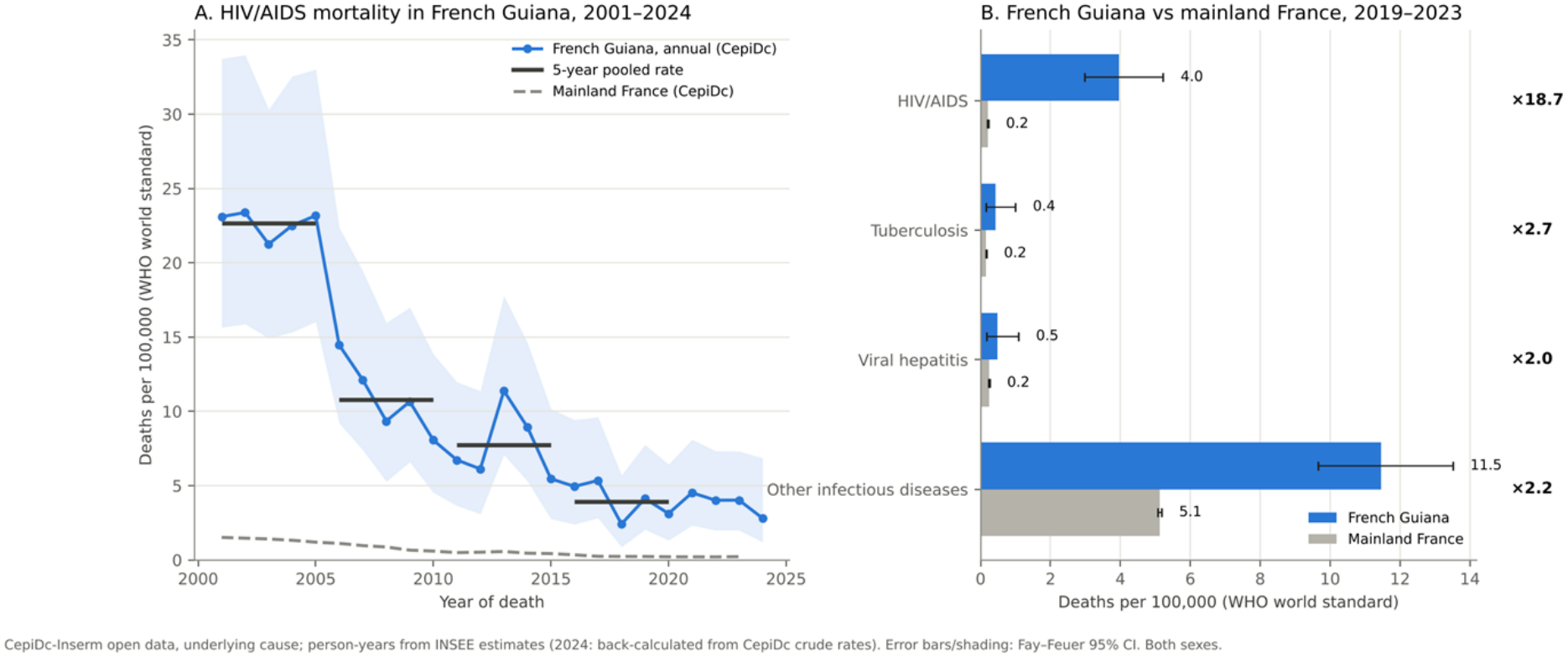
(A) Annual and 5-year pooled AIDS mortality in French Guiana and mainland France, both sexes. (B) Infectious causes of death, French Guiana vs mainland France, 2019–2023, with rate ratios (WHO world standard; Fay–Feuer 95% CI). CépiDc-Inserm open data; mainland counts rebuilt from crude rates and INSEE populations.

Mainland France fell just as fast, from 1.38 to 0.21 per 100,000. The ratio therefore did not shrink: French Guiana was 16 times the mainland rate in 2001–2005 and 19 times in 2019–2023. AIDS was by far the largest excess in the infectious-disease panel; tuberculosis (×2.7), viral hepatitis (×2.0) and other infectious diseases (×2.2) were also higher (Figure 2B).

Deaths were concentrated in working age. In 2015–2024, 65 of 103 AIDS deaths (63%) occurred before age 55, with rates of 7.7–8.3 per 100,000 at 35–64 years, against 0.3–0.8 in mainland France.

### Men and women

In men, the rate fell from 29.5 to 5.4 per 100,000 (−9.9% a year); in women, from 15.8 to 2.8 (−9.2% a year). Men’s rates were 1.3–1.9 times women’s. In mainland France the male excess was larger (men 0.31, women 0.12 in 2019–2023), so the ratio to the mainland was higher in women: ×25 in 2001–2005 and ×23 in 2019–2023, against ×14 and ×17 in men (Figure 5B).

### French Guiana in the GBD frame

GBD put mainland France’s HIV/AIDS death rate at 0.45 per 100,000 in 2019–2023, 2.1 times the CépiDc rate (k = 2.12 (1.79–2.44)). Scaled by k, French Guiana’s observed rate of 4.0 became a GBD-equivalent rate of 8.3 (6.0–11.2) per 100,000, 19 (14–24) times France, or about 22 deaths a year on the GBD scale against about 10 certified (Table 1).

This placed French Guiana level with Amazonas and Pará (it exceeded them in 41% and 41% of draws), above Maranhão (75%), Roraima (77%) and Amapá (84%), above Mato Grosso (97%), and above Venezuela, Peru, Bolivia, Colombia, Ecuador, Rondônia, Tocantins and Acre in more than 98% of draws. It never exceeded Suriname or Guyana, whose rates were about twice as high (Figures 3 and 4). YLLs were 400 (285–544) per 100,000 and YLDs 97 (58–149), for DALYs of 499 (377–652). The YLD share (19%) was higher than in the GBD locations, reflecting the large number of people living with HIV on treatment.

**Figure 3.**
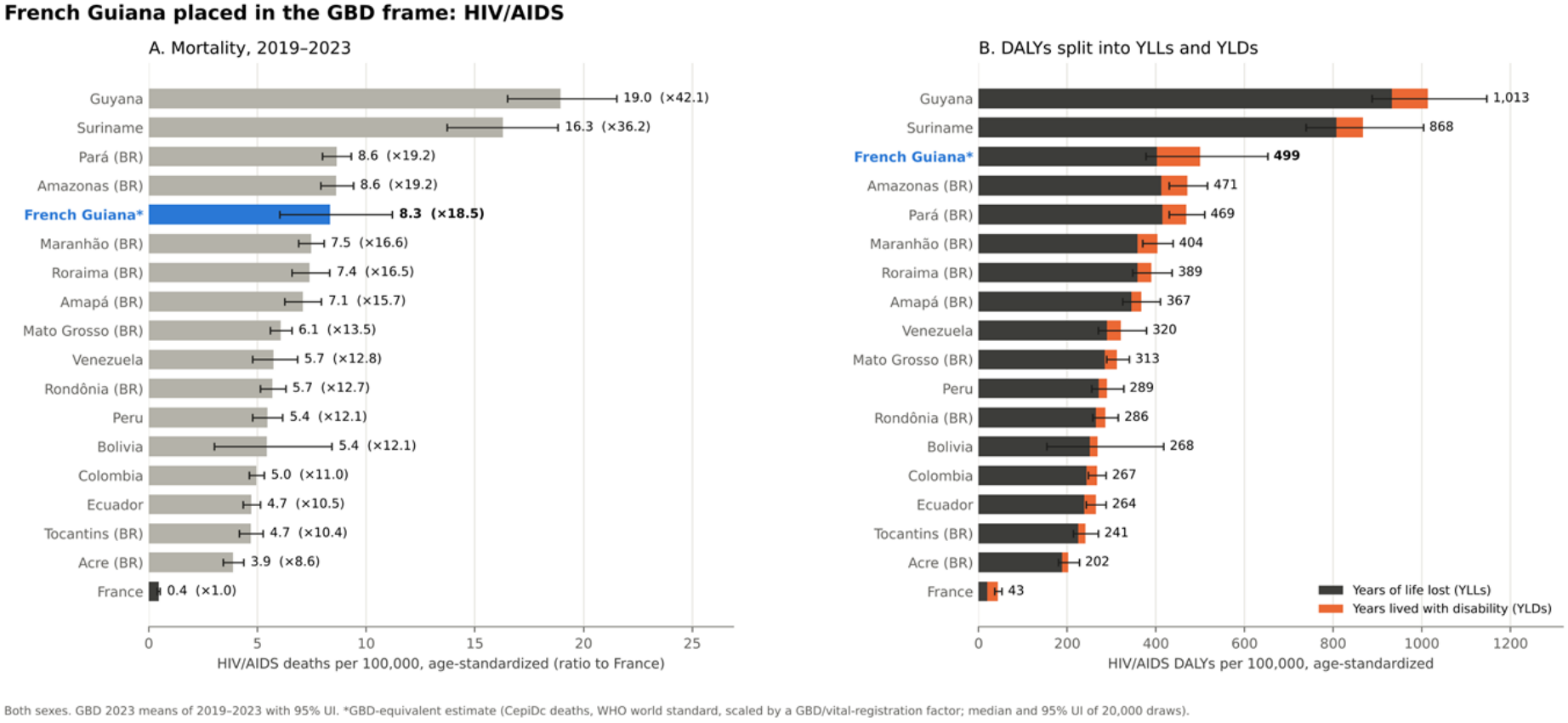
Age-standardized HIV/AIDS deaths (A, with ratio to France) and DALYs split into YLLs and YLDs (B), with 95% UIs. GBD locations: 2019–2023 means; *French Guiana: GBD-equivalent estimate, 2019–2023, calibrated on mainland France CépiDc data.

**Figure 4.**
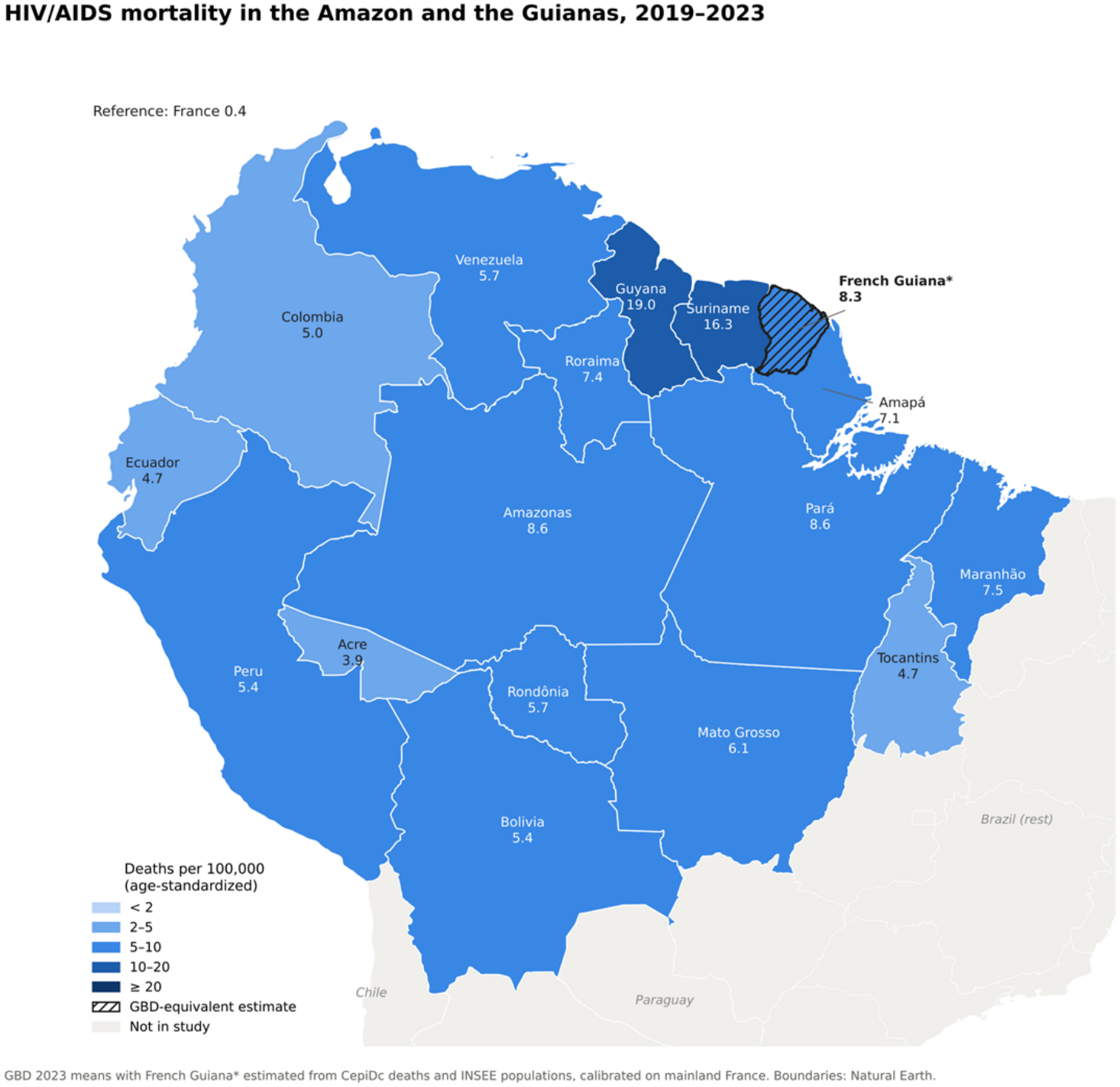
Age-standardized HIV/AIDS deaths per 100,000 by country or Brazilian state (GBD 2023 means, 2019–2023). *French Guiana (hatched): GBD-equivalent estimate, calibrated on mainland France. Boundaries: Natural Earth.

### Trajectories

Set against GBD estimates over time, French Guiana followed a different path from its neighbors (Figure 5A). In 2001–2005 its observed rate alone was below Guyana and Suriname but three to five times the Brazilian Amazon states. Since then it has fallen while the Brazilian Amazon states have stayed flat, so that by 2019–2023 French Guiana, on the GBD scale, had joined them.

**Figure 5.**
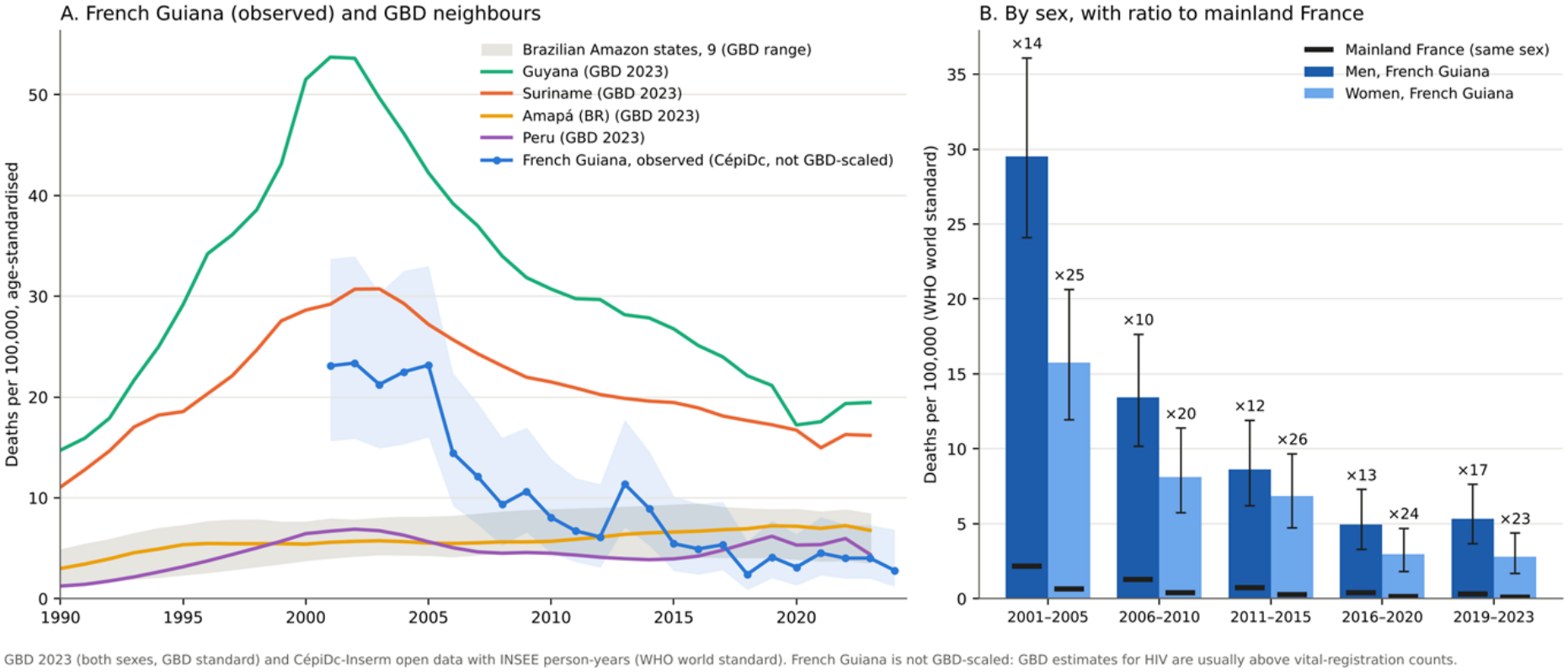
(A) Observed AIDS mortality in French Guiana (CépiDc, not GBD-scaled) against GBD 2023 estimates for Guyana, Suriname, Amapá, Peru and the range of the nine Brazilian Legal Amazon states. (B) AIDS mortality in French Guiana by sex and period, with mainland France for the same sex and the rate ratio.

### Men and women on the GBD scale

With sex-specific calibration (k = 2.03 in men and 2.29 in women), the GBD-equivalent death rate was 10.7 (7.0–15.7) per 100,000 in men, 17 (12–23) times French men, and 6.3 (3.8–9.8) in women, 23 (14–35) times French women; about 13 and 9 deaths a year on the GBD scale (Figure 6).

**Figure 6.**
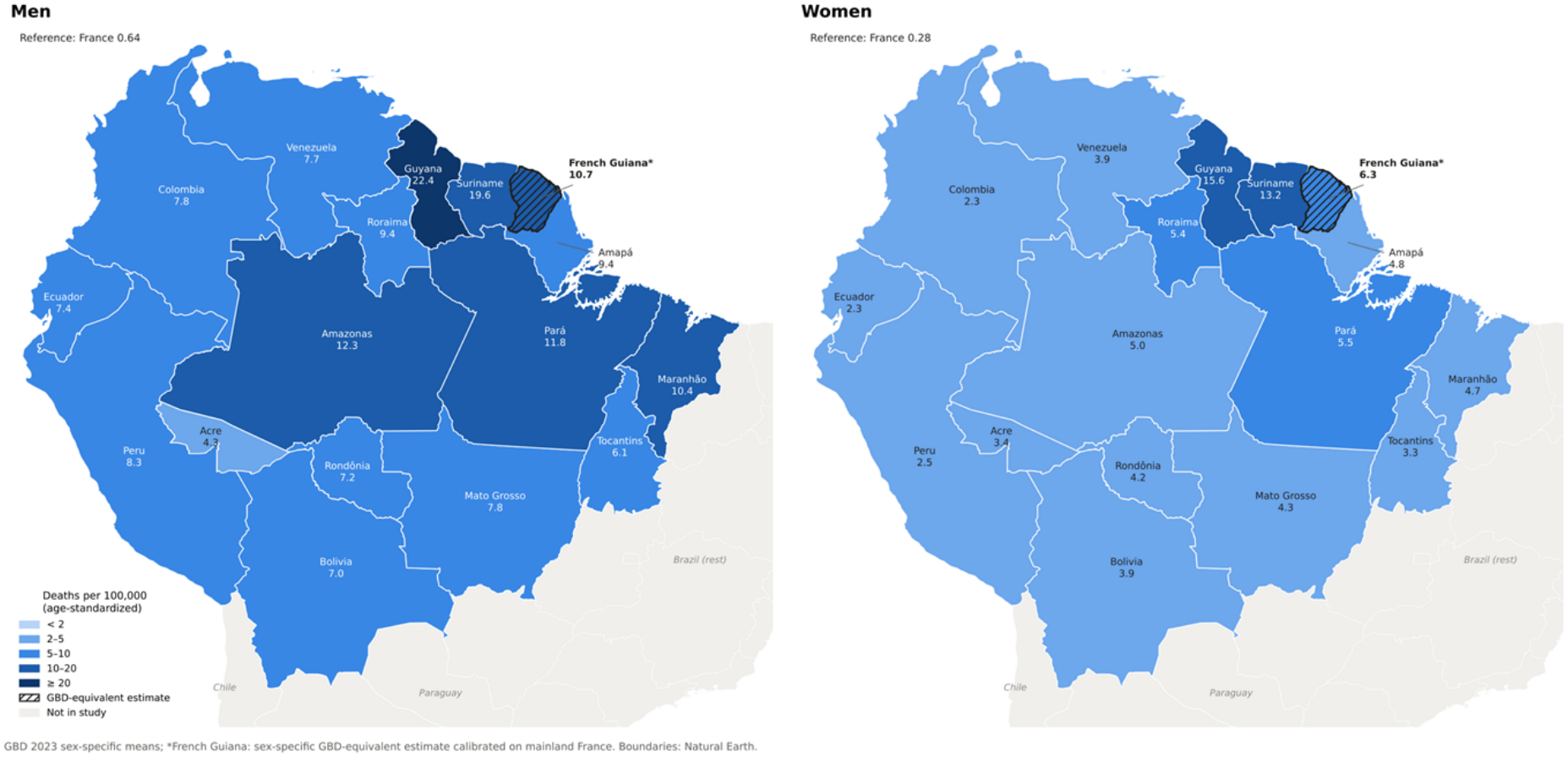
Age-standardized HIV/AIDS deaths per 100,000 in men (left) and women (right), GBD 2023 means 2019–2023. *French Guiana (hatched): sex-specific GBD-equivalent estimate, calibrated on mainland France. Note the different reference rates for France.

The two sexes did not rank the same way among the neighbors. French Guianese men were level with Pará and Amazonas (they exceeded them in 31% and 24% of draws), where male mortality is high. French Guianese women exceeded Amazonas in 82% of draws, Pará in 70% and Amapá in 85%, and ranked third in the region after Guyana (15.6) and Suriname (13.2). Neither sex reached the level of Suriname or Guyana. French Guianese men also exceeded Maranhão in 55% of draws, Mato Grosso in 93% and Peru in 88%; women exceeded Maranhão in 88%, Mato Grosso in 93% and Peru in more than 99%. DALYs were 622 (442–861) per 100,000 in men and 391 (249–595) in women.

### Sex patterns across territories

The sex balance of the epidemic differed sharply across the region (Figure 7A, Table 2). In Suriname, Acre, Rondônia, Guyana, Mato Grosso, Tocantins and Roraima, the male:female prevalence ratio was 0.87–1.25: as many women as men, or more, were living with HIV. In Suriname, new infections were more frequent in women than in men (ratio 0.71). At the other end, Ecuador, Bolivia, Amazonas, France, Venezuela, Colombia and Peru had male:female prevalence ratios of 2.4–3.5, the pattern of epidemics concentrated in men who have sex with men. Maranhão, Amapá and Pará were in between (1.5–1.7).

**Table 2.**
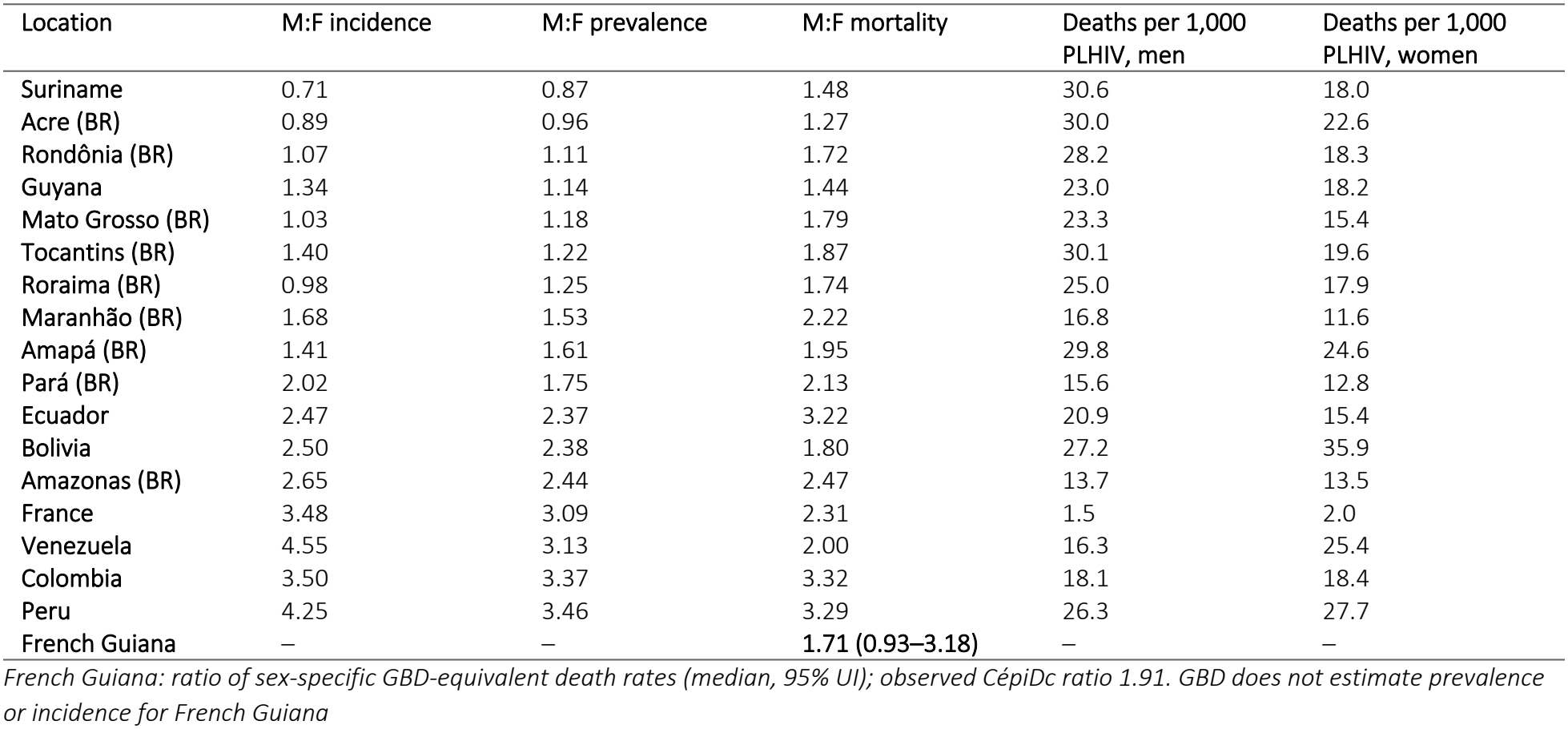
Male:female (M:F) ratios of age-standardized HIV incidence, prevalence and mortality, and deaths per 1,000 people living with HIV (PLHIV), 2019–2023 (GBD 2023).

| Location | M:F incidence | M:F prevalence | M:F mortality | Deaths per 1,000 PLHIV, men | Deaths per 1,000 PLHIV, women |
|---|---|---|---|---|---|
| Suriname | 0.71 | 0.87 | 1.48 | 30.6 | 18.0 |
| Acre (BR) | 0.89 | 0.96 | 1.27 | 30.0 | 22.6 |
| Rondônia (BR) | 1.07 | 1.11 | 1.72 | 28.2 | 18.3 |
| Guyana | 1.34 | 1.14 | 1.44 | 23.0 | 18.2 |
| Mato Grosso (BR) | 1.03 | 1.18 | 1.79 | 23.3 | 15.4 |
| Tocantins (BR) | 1.40 | 1.22 | 1.87 | 30.1 | 19.6 |
| Roraima (BR) | 0.98 | 1.25 | 1.74 | 25.0 | 17.9 |
| Maranhão (BR) | 1.68 | 1.53 | 2.22 | 16.8 | 11.6 |
| Amapá (BR) | 1.41 | 1.61 | 1.95 | 29.8 | 24.6 |
| Pará (BR) | 2.02 | 1.75 | 2.13 | 15.6 | 12.8 |
| Ecuador | 2.47 | 2.37 | 3.22 | 20.9 | 15.4 |
| Bolivia | 2.50 | 2.38 | 1.80 | 27.2 | 35.9 |
| Amazonas (BR) | 2.65 | 2.44 | 2.47 | 13.7 | 13.5 |
| France | 3.48 | 3.09 | 2.31 | 1.5 | 2.0 |
| Venezuela | 4.55 | 3.13 | 2.00 | 16.3 | 25.4 |
| Colombia | 3.50 | 3.37 | 3.32 | 18.1 | 18.4 |
| Peru | 4.25 | 3.46 | 3.29 | 26.3 | 27.7 |
| French Guiana | – | – | 1.71 (0.93–3.18) | – | – |

*French Guiana: ratio of sex-specific GBD-equivalent death rates (median, 95% UI); observed CépiDc ratio 1.91. GBD does not estimate prevalence or incidence for French Guiana*

**Figure 7.**
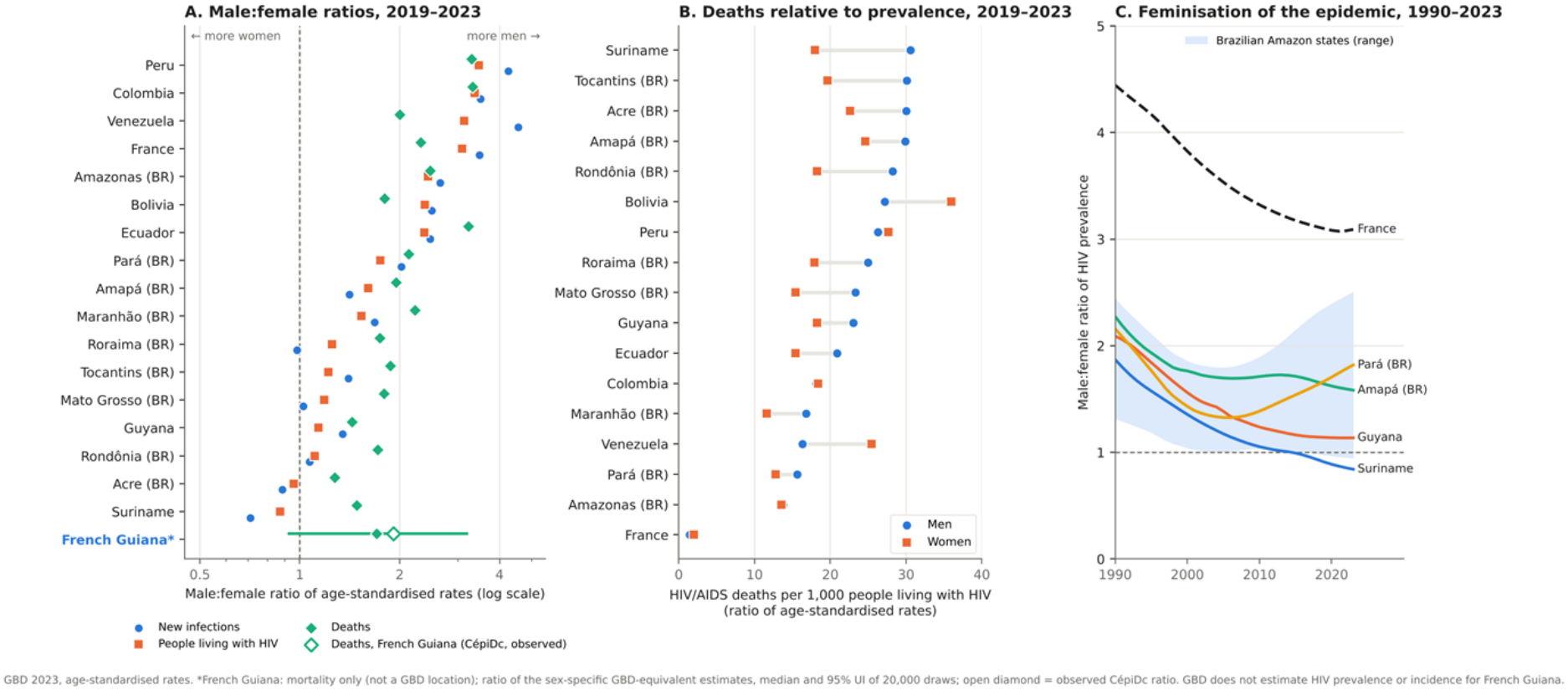
Sex differences in HIV across the Amazon and the Guianas (GBD 2023, age-standardized). (A) Male:female ratios of incidence, prevalence and mortality, 2019–2023; French Guiana: mortality only, GBD-equivalent (median, 95% UI) and observed CépiDc ratio. (B) Deaths per 1,000 people living with HIV by sex. (C) Male:female prevalence ratio, 1990–2023.

Mortality was more male-biased than prevalence almost everywhere in the generalised epidemics: in Suriname the male:female ratio was 0.87 for prevalence but 1.48 for deaths. Men living with HIV died more often than women: 31 against 18 deaths per 1,000 people living with HIV in Suriname, 28 against 18 in Rondônia and 23 against 18 in Guyana (Figure 7B). Where women were a minority of people living with HIV, the gap reversed: 25 against 16 in Venezuela, 36 against 27 in Bolivia, 28 against 26 in Peru, and 2.0 against 1.5 in France.

French Guiana’s male:female mortality ratio was 1.71 (95% UI 0.93–3.18) on the GBD scale and 1.91 in the observed CépiDc data. This is close to Guyana (1.44), Suriname (1.48) and Roraima (1.74), and below France (2.31), Colombia, Ecuador and Peru (above 3), although its UI is wide.

Over time the epidemic became more female across the Guiana Shield (Figure 7C). The male:female prevalence ratio fell from 1.87 in 1990 to 0.84 in 2023 in Suriname and from 2.09 to 1.14 in Guyana. It fell in France too, from 4.4 to 3.1, but in 2023 was still about three times that of the Guianas. In Pará and Amazonas the trend reversed in the 2000s, with the ratio rising back to 1.8 and 2.5.

### Prevalence and incidence

GBD placed HIV prevalence in 2019–2023 highest in Guyana (916 per 100,000, age-standardized) and Suriname (688), followed by Amazonas (633), Pará (593) and Maranhão (514); the other locations were at 148–339, against 288 in France (Figure 8A). New infections ranked differently: they were most frequent in Amazonas (51 per 100,000 a year), Pará (46), Suriname (44), Guyana (39) and Maranhão (38), against 9–24 elsewhere and 14 in France (Figure 8B). New infections relative to prevalence were higher in Amazonas, Pará and Maranhão (7–8%) than in Guyana (4%), consistent with epidemics that are still growing.

**Figure 8.**
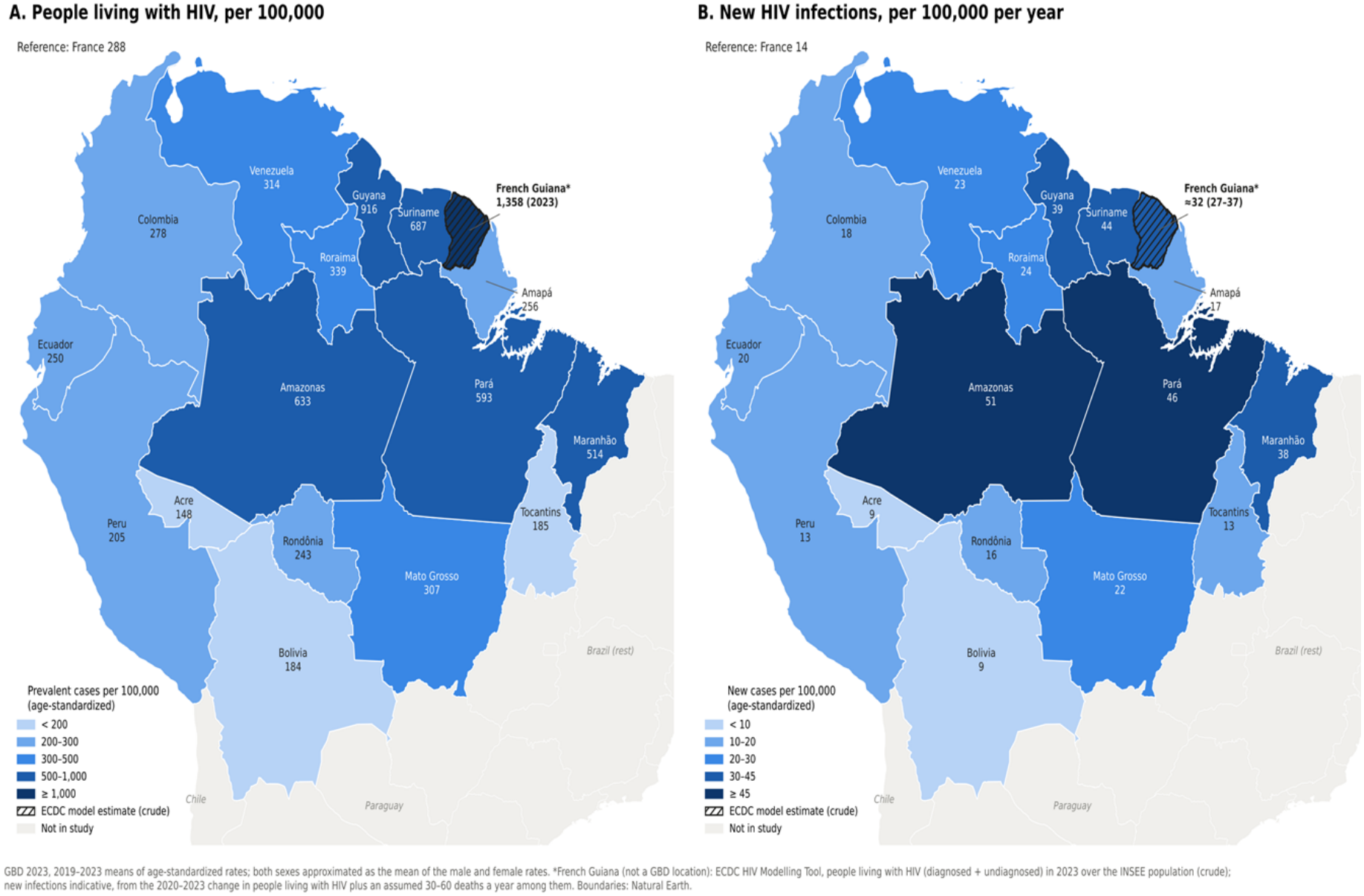
HIV prevalence (A) and new HIV infections (B) per 100,000, 2019–2023 (GBD 2023, age-standardized; both sexes approximated as the mean of the male and female rates). *French Guiana (hatched): people living with HIV in 2023 from the ECDC HIV Modelling Tool over the INSEE population; new infections indicative.

For French Guiana, the ECDC HIV Modelling Tool estimated about 4,090 people living with HIV in 2023, of whom 3,880 were diagnosed and 210 undiagnosed (95% diagnosed). This is a crude prevalence of 1,358 per 100,000 (1.4%), above every GBD location; age-standardization to the world population, which has a larger share of adults aged 25–64 than French Guiana, would probably raise rather than lower it. The number of people living with HIV grew by about 50 a year in 2020–2023; with 30–60 deaths a year among them, this suggests about 80–110 new infections a year, an indicative incidence of 27–37 per 100,000, close to Guyana and Suriname and below Amazonas and Pará. New infections relative to prevalence (about 2.4%) were the lowest in the region apart from France.

### Simulated histoplasmosis-attributable deaths

Applying a common histoplasmosis-attributable fraction of 20–40% to HIV/AIDS mortality yielded substantial geographical differences across the Amazon and the Guianas. The highest simulated age-standardized mortality rates were observed in Guyana (5.7 [95% UI 4.1–7.5] per 100,000) and Suriname (4.9 [3.5–6.5]), followed by Pará and Amazonas (both 2.6 [1.9–3.3]) (Figure 9). These estimates represent exploratory scenarios based on historical evidence and do not account for differences in the evolution of histoplasmosis diagnosis and treatment.

**Figure 9.**
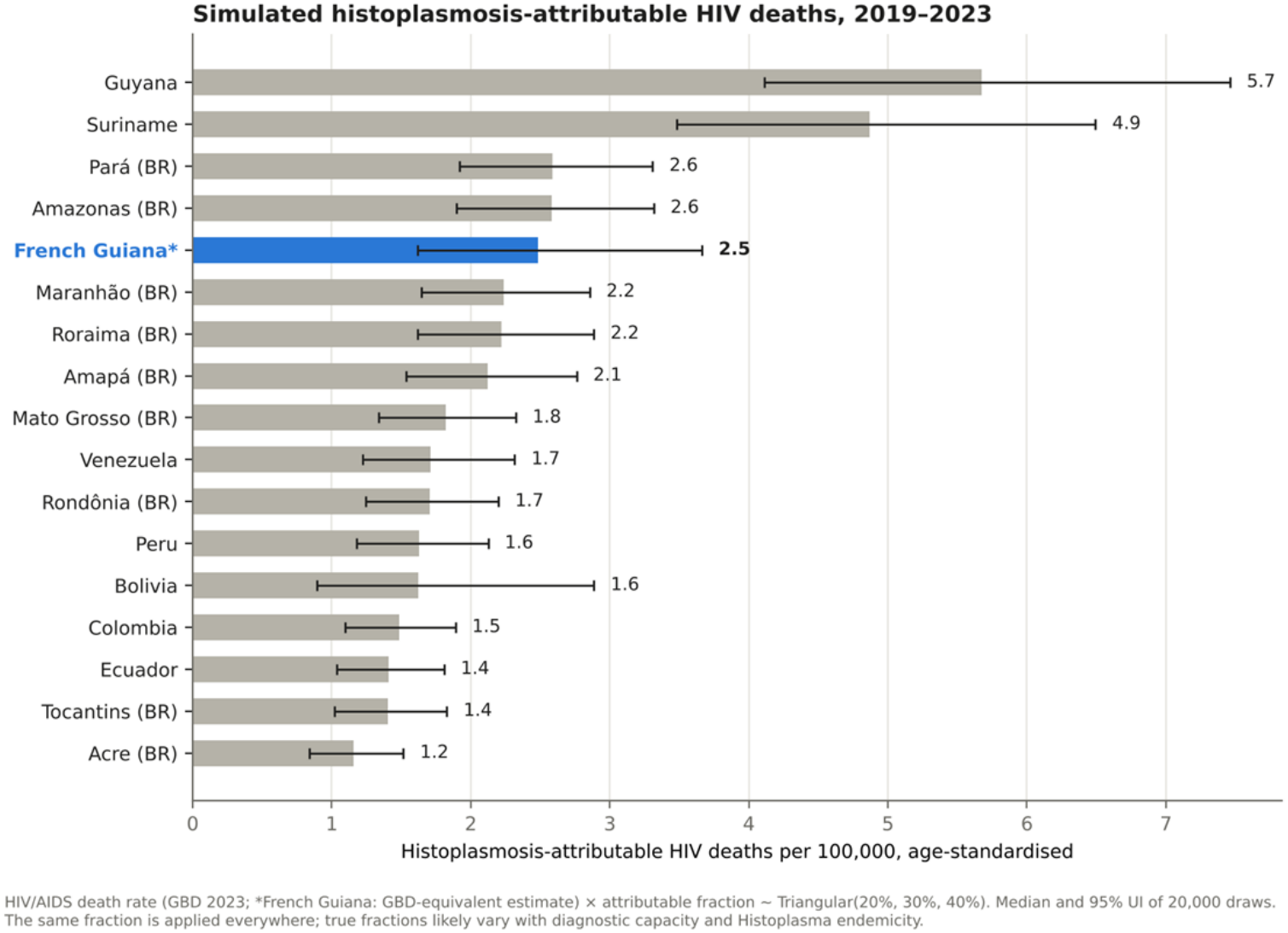
Exploratory simulation of histoplasmosis-attributable HIV mortality per 100,000 population (age-standardized), 2019–2023. Regional estimates were calculated by multiplying HIV/AIDS mortality (GBD 2023; French Guiana: GBD-equivalent estimate) by an attributable fraction drawn from Triangular(20%, 30%, 40%). *Bars represent medians and error bars 95% uncertainty intervals from 20,000 simulations. The French Guiana estimate shown (2.5 per 100,000) represents the common-fraction scenario rather than its locally informed estimate. A separate calculation using approximately 16 diagnosed histoplasmosis episodes annually and 3.9% early case fatality yielded 0.62 expected early deaths annually (crude rate 0.21 per 100,000). The two approaches estimate different outcomes and should not be interpreted as directly comparable measures of contemporary mortality*.

French Guiana warranted separate consideration because of its longstanding clinical experience in diagnosing and treating HIV-associated histoplasmosis. Applying the common regional attributable fraction to its GBD-equivalent HIV mortality yielded 2.5 (1.6–3.7) deaths per 100,000, corresponding to approximately 6.6 (4.3–9.8) deaths annually. However, local longitudinal data documented approximately 16 diagnosed histoplasmosis episodes annually during 2012–2019, with early case fatality declining to 3.9%. Applying this case-fatality proportion to the historical annual episode count yielded approximately 0.62 early deaths annually, equivalent to a crude rate of 0.21 per 100,000 inhabitants. At hypothetical case-fatality proportions of 2%, 3%, 5% and 10%, expected early deaths ranged from 0.32 to 1.60 annually. These locally informed estimates concern mortality following diagnosed histoplasmosis and are not directly equivalent to the fraction of all HIV deaths attributable to histoplasmosis.

For Suriname, the historical hospital-based Guiana Shield cohort reported 30-day case fatality of 17.5%. Applying this proportion to hypothetical annual counts of 50, 100 and 150 histoplasmosis episodes would yield approximately 9, 18 and 26 early deaths annually, respectively. Because the published case-fatality estimate was derived from a combined French Guiana–Suriname cohort and reliable contemporary national episode counts were unavailable, these figures were considered sensitivity scenarios rather than estimates of current national mortality.

Under the common regional attributable-fraction model, approximately 701 (520–884) histoplasmosis-attributable HIV deaths annually were simulated across the three Guianas and nine Brazilian Legal Amazon states, including 83 in the Guiana Shield and 618 in the Brazilian Amazon. Pará (210), Maranhão (152) and Amazonas (102) contributed the largest absolute numbers. Including Colombia, Venezuela, Peru, Ecuador and Bolivia as whole countries increased the simulated total to 2,976 (2,202–3,778) annually. The lower and upper attributable-fraction scenarios yielded 1,487 and 3,727 deaths, respectively (Table 3). These totals describe the original common-fraction model and should not be interpreted as locally calibrated estimates of contemporary histoplasmosis mortality.

**Table 3.** Exploratory estimates of annual HIV/AIDS deaths, simulated histoplasmosis-attributable mortality, and locally informed early-mortality scenarios in the Amazon and the Guianas, 2019–2023.

| Location | Population (millions) | HIV/AIDS deaths per year (95% UI) | Histoplasmosis deaths per year, main scenario (95% UI) | Low scenario | High scenario |
|---|---|---|---|---|---|
| French Guiana | 0.29 | 22 (16–30) | 7 (4–10) | 3 | 8 |
| Suriname | 0.62 | 101 (86–118) | 30 (22–40) | 15 | 38 |
| Guyana | 0.81 | 153 (134–175) | 46 (33–60) | 23 | 58 |
| Guiana Shield | 1.72 | 277 (251–306) | **83 (61–107)** | 42 | 104 |
| Acre (BR) | 0.83 | 32 (28–36) | 10 (7–13) | 5 | 12 |
| Amapá (BR) | 0.73 | 52 (46–58) | 15 (11–20) | 8 | 19 |
| Amazonas (BR) | 3.94 | 340 (311–370) | 102 (75–130) | 51 | 127 |
| Pará (BR) | 8.12 | 701 (649–757) | 210 (154–268) | 105 | 263 |
| Rondônia (BR) | 1.58 | 90 (81–100) | 27 (20–35) | 13 | 34 |
| Roraima (BR) | 0.64 | 47 (42–53) | 14 (10–18) | 7 | 18 |
| Tocantins (BR) | 1.51 | 71 (63–79) | 21 (15–27) | 11 | 27 |
| Maranhão (BR) | 6.78 | 506 (469–547) | 152 (112–194) | 76 | 190 |
| Mato Grosso (BR) | 3.66 | 222 (205–241) | 66 (49–85) | 33 | 83 |
| Brazilian Amazon states | 27.79 | 2,062 (1,987–2,140) | **618 (458–780)** | 309 | 774 |
| Guianas + Brazilian Amazon | 29.51 | 2,340 (2,261–2,423) | **701 (520–884)** | 351 | 879 |
| Colombia | 52.10 | 2,583 (2,410–2,768) | 774 (572–986) | 387 | 969 |
| Venezuela | 28.30 | 1,624 (1,354–1,946) | 485 (344–658) | 242 | 607 |
| Peru | 33.70 | 1,836 (1,618–2,083) | 549 (398–719) | 274 | 687 |
| Ecuador | 18.00 | 848 (778–922) | 254 (187–326) | 127 | 318 |
| Bolivia | 12.40 | 672 (400–1,126) | 200 (111–357) | 99 | 250 |
| Colombia, Venezuela, Peru, Ecuador, Bolivia (whole countries) | 144.50 | 7,584 (7,074–8,196) | **2,273 (1,680–2,906)** | 1,136 | 2,848 |
| All study locations | 174.01 | 9,927 (9,404–10,540) | **2,976 (2,202–3,778)** | 1,487 | 3,727 |

Notes: Regional histoplasmosis-attributable deaths were simulated by multiplying age-standardized HIV/AIDS mortality rates (GBD 2023; French Guiana: GBD-equivalent estimate) by an assumed attributable fraction: main scenario Triangular(20%, 30%, 40%), low scenario Uniform(10%, 20%), and high scenario Uniform(30%, 45%). Absolute death counts were approximated by multiplying age-standardized rates by population size and may be biased in populations with different age structures. For French Guiana, an alternative locally informed calculation used approximately 16 diagnosed histoplasmosis episodes annually during 2012–2019 and 3.9% early case fatality, yielding 0.62 expected early deaths annually. For Suriname, the historical 17.5% case-fatality proportion was derived from a combined French Guiana–Suriname hospital cohort; applying it to hypothetical annual episode counts of 50–150 yields approximately 9–26 deaths. Neither calculation represents a validated contemporary national mortality estimate. Locally informed estimates and regional attributable-fraction simulations measure different outcomes and should not be combined into a single regional total. Colombia, Venezuela, Peru, Ecuador and Bolivia were included as whole countries rather than Amazonian subnational populations. Population sources: IBGE 2022 census and national population estimates.

## Discussion

The progression of each territory toward the UNAIDS 95-95-95 targets presumably explains much of the mortality gradients we observed. In French Guiana, modelling estimates suggest that about 95% of people living with HIV are diagnosed, and the three targets were reported as reached in 2025.[7] This was achieved despite the territory having the highest prevalence in the region. AIDS mortality fell by more than 80% in two decades, faster than in Suriname and Guyana and unlike the Brazilian Amazon, where GBD shows little or no decline (at most about 20% from the peak, in Maranhão and Mato Grosso). On the GBD scale, French Guiana moved from the high-mortality group of the Guiana Shield to the level of Amazonas and Pará, and half the rate of Suriname and Guyana. Brazil as a whole reported that 93% of people living with HIV were diagnosed, 81% of those diagnosed were on antiretroviral treatment, and 96% of those treated were virally suppressed in 2024. [19] In the Norte region, 15.6% of people linked to care were lost to follow-up and viral suppression was the lowest of the five regions. Peru reported 91% diagnosed and 91% treated, but no national viral suppression estimate.[20] The Guiana Shield countries appear furthest from the targets. In Suriname, the most recent UNAIDS estimates put the cascade at 50-71-89[21], and a national cohort found that only 63% of those diagnosed initiated treatment.[22] Guyana has announced that 95% of people living with HIV know their status, but recent treatment and suppression figures are lacking. Across Latin America, around 14% of people living with HIV remain undiagnosed, and incidence outside Brazil has stagnated or risen.[23] Comparable cascades are not available for the Amazonian subnational units of Colombia, Ecuador, Bolivia, Venezuela or Peru, so national figures probably hide worse situations in remote Amazonian areas. There, distance, transport costs, mobility linked to gold mining and cross-border movement, and stigma all erode continuity of care. French Guiana combines high HIV prevalence with relatively high treatment coverage and a substantial decline in AIDS mortality. Although the contribution of individual interventions cannot be determined from these data, the findings are consistent with a beneficial role of broad access to antiretroviral treatment and HIV care. It also emphasizes that the remaining gap, the undiagnosed and those lost to follow-up, is where most avoidable deaths occur.

The sex patterns in French Guiana are similar as in the Guiana Shield. Across the region, two kinds of epidemic coexist: gender-balanced, heterosexual epidemics in Suriname, Guyana and several Brazilian Amazon states, in which women now represent half or more of people living with HIV, and male-predominant epidemics in France, Colombia, Venezuela, Ecuador and Amazonas.

The steep decline in AIDS mortality in French Guiana from 2006 onwards coincided with the expansion of antiretroviral treatment, HIV testing, and improvements in the diagnosis and management of opportunistic infections, particularly disseminated histoplasmosis. [9,11] Histoplasmosis has historically been a leading AIDS-defining infection and cause of death in French Guiana, where decades of clinical experience have contributed to greater awareness, improved diagnostic capacity and earlier antifungal treatment. Applying the 3.9% French Guianese case-fatality proportion to the historical mean of approximately 16 diagnosed episodes annually yielded fewer than one expected early death per year, considerably below the estimate obtained using the common regional attributable fraction. This does not account for undiagnosed histoplasmosis or later attributable deaths, but suggests that extrapolating historical regional mortality assumptions to contemporary French Guiana may substantially overestimate mortality following diagnosed histoplasmosis. The more rapid decline in AIDS mortality in French Guiana than in Suriname and Guyana, despite shared viral lineages and population movements, is consistent with a beneficial role of broad access to antiretroviral treatment, opportunistic infection diagnostics and specialized care, although their respective contributions cannot be established from these ecological data.[24]

The persistent excess may reflect late diagnosis with too many persons still diagnosed at a very advanced stage, and this share has not fallen [13]. Another problem is the hidden reservoir of infected persons unaware of their infection as reflected by the fact that the 95*95*95 milestone has not been reached in much of the Amazon region. In French Guiana, by contrast, the ECDC model estimates the undiagnosed share at about 5% in 2023, yet prevalence is the highest in the region and new infections continue at a rate close to that of Guyana and Suriname. Finally, loss to follow-up remains a problem, which is frequent among mobile and migrant patients, but is also the result of stigma and discrimination.[25]

Across the region, wherever the epidemic is gender-balanced, men living with HIV died more often than women. This is consistent with later diagnosis and poorer retention in men, whereas women are tested routinely in antenatal care. In French Guiana, antenatal screening has been near-universal for decades, and men are over-represented among late presenters.[26] Where women are a minority of people living with HIV, as in Venezuela, Bolivia and France, the opposite is observed, which may reflect lower suspicion of HIV in women and later diagnosis.

This study is the first to place French Guiana on the GBD scale for HIV/AIDS, but several limitations warrant consideration. First, calibration assumes that the discrepancy between GBD estimates and certified causes of death is similar in French Guiana and mainland France; differences in death certification could bias the estimates. YLDs were also extrapolated from mainland France using assumed prevalence ratios. Second, histoplasmosis-attributable mortality was simulated using historical fractions that may no longer reflect improvements in diagnosis and treatment, particularly in French Guiana, where early case fatality among diagnosed patients has fallen to 3.9%. The locally informed scenario yielded substantially fewer early deaths, but this measures a different outcome from total histoplasmosis-attributable HIV mortality, which may include undiagnosed infections. Similarly, the 17.5% case-fatality assumption for Suriname came from a combined Guiana Shield hospital cohort. These simulations should therefore be regarded as illustrative rather than precise contemporary estimates. Third, the small number of annual AIDS deaths in French Guiana produces unstable rates, while estimates for several neighboring countries rely partly on modelling and incomplete vital registration. Finally, for Colombia, Venezuela, Peru, Ecuador and Bolivia, only national GBD estimates were available. Because much of their population lives outside Amazonia, these estimates may not adequately represent the burden in remote Amazonian areas, where access to care is generally more difficult.

In conclusion, AIDS mortality in French Guiana declined substantially between 2001 and 2024 and is now comparable to that of several Brazilian Amazonian states, although it remains approximately twenty times higher than in mainland France. This decline occurred alongside expanded access to antiretroviral treatment, improved HIV care and major advances in the diagnosis and management of opportunistic infections, particularly disseminated histoplasmosis. The contrast with neighboring territories highlights the potential importance of healthcare access and specialized clinical expertise, while also emphasizing the limitations of applying common mortality assumptions across settings with different diagnostic and therapeutic capacities. Earlier HIV diagnosis, sustained retention in care and continued progress toward the UNAIDS 95-95-95 targets remain essential to reducing avoidable AIDS mortality across the region.

## Data Availability

The data is publicly available at https://www.healthdata.org/ and https://opendata-cepidc.inserm.fr/

https://www.healthdata.org/

## Notes

### Competing Interest Statement

The authors have declared no competing interest.

### Clinical Trial

NA

### Author Declarations

The present study does not require ethical clearance

## References

1. Bello G, Nacher M, Divino F, Darcissac E, Mir D, Lacoste V. The HIV-1 Subtype B Epidemic in French Guiana and Suriname Is Driven by Ongoing Transmissions of Pandemic and Non-pandemic Lineages. Front Microbiol. 2018;9:1738. doi:10.3389/fmicb.2018.01738

2. Arantes I, Bello G, Darcissac E, Lacoste V, Nacher M. Using phylogenetic surveillance and epidemiological data to understand the HIV-1 transmission dynamics in French Guiana. AIDS. 2021;35:979–984. doi:10.1097/QAD.0000000000002817

3. Divino F, de Lima Guerra Corado A, Gomes Naveca F, Stefani MMA, Bello G. High Prevalence and Onward Transmission of Non-Pandemic HIV-1 Subtype B Clades in Northern and Northeastern Brazilian Regions. PLoS One. 2016;11:e0162112. doi:10.1371/journal.pone.0162112

4. Nacher M, Adriouch L, Van Melle A, Parriault M-C, Adenis A, Couppié P. Country of infection among HIV-infected patients born abroad living in French Guiana. PLoS One. 2018;13:e0192564. doi:10.1371/journal.pone.0192564

5. Nacher M, Vantilcke V, Parriault MC, Van Melle A, Hanf M, Labadie G, et al. What is driving the HIV epidemic in French Guiana? Int J STD AIDS. 2010;21:359–361. doi:10.1258/ijsa.2010.009570

6. Nacher M, Adriouch L, Huber F, Vantilcke V, Djossou F, Elenga N, et al. Modeling of the HIV epidemic and continuum of care in French Guiana. PLoS One. 2018;13:e0197990. doi:10.1371/journal.pone.0197990

7. Nacher M, Lucarelli A, Vergeade F, Rabier S, Delin C, Epelboin L, et al. Evolution of HIV epidemic trends and diagnostic delays in French Guiana: how can we make further progress? BMC Infect Dis. 2026;26:1612. doi:10.1186/s12879-026-12851-4

8. Nacher M, Huber F, Adriouch L, Djossou F, Adenis A, Couppié P. Temporal trend of the proportion of patients presenting with advanced HIV in French Guiana: stuck on the asymptote? BMC Res Notes. 2018;11:831. doi:10.1186/s13104-018-3944-y

9. Adenis A, Nacher M, Hanf M, Vantilcke V, Boukhari R, Blachet D, et al. HIV-associated histoplasmosis early mortality and incidence trends: from neglect to priority. PLoS Negl Trop Dis. 2014;8:e3100. doi:10.1371/journal.pntd.0003100

10. Nacher M, Marshall E, Bani-Sadr F, Peugny S, Denis B, Ouedraogo E, et al. Disseminated Histoplasmosis in Persons Living with HIV, France and Overseas Territories,1992–2021 - Volume 31, Number 7—July 2025 - Emerging Infectious Diseases journal - CDC. [cited 11 Nov 2026]. doi:10.3201/eid3107.241931

11. Nacher M, Adenis A, Guarmit B, Lucarelli A, Blanchet D, Demar M, et al. What is AIDS in the Amazon and the Guianas in the 90-90-90 era? PLoS One. 2020;15:e0236368. doi:10.1371/journal.pone.0236368

12. Nacher M, Adenis A, Adriouch L, Dufour J, Papot E, Hanf M, et al. What is AIDS in the Amazon and the Guianas? Establishing the burden of disseminated histoplasmosis. Am J Trop Med Hyg. 2011;84:239–240. doi:10.4269/ajtmh.2011.10-0251

13. AIDSinfo | UNAIDS. [cited 20 Aug 2024]. Available: https://aidsinfo.unaids.org/

14. GBD Compare. In: Institute for Health Metrics and Evaluation [Internet]. [cited 11 Nov 2026]. Available: https://vizhub.healthdata.org/gbd-compare

15. OpenData CépiDc. [cited 11 Nov 2026]. Available: https://opendata-cepidc.inserm.fr/

16. Adenis AA, Valdes A, Cropet C, McCotter OZ, Derado G, Couppie P, et al. Burden of HIV-associated histoplasmosis compared with tuberculosis in Latin America: a modelling study. Lancet Infect Dis. 2018;18:1150–1159. doi:10.1016/S1473-3099(18)30354-2

17. Rakislova N, Hurtado JC, Palhares AEM, Ferreira L, Freire M, Lacerda M, et al. High prevalence and mortality due to Histoplasma capsulatum in the Brazilian Amazon: An autopsy study. PLoS Negl Trop Dis. 2021;15:e0009286. doi:10.1371/journal.pntd.0009286

18. Françoise U, Nacher M, Donald SM, Van Eer M, Scheel C, Chiller T, et al. Prevalence of HIV-associated histoplasmosis in the Guiana Shield: A prospective study. International Journal of Infectious Diseases. 2026;169:108795. doi:10.1016/j.ijid.2026.108795

19. MONITORAMENTO INTEGRADO DO CUIDADO DO HIV E DA AIDS - 2025. [cited 11 Nov 2026]. Available: https://www.gov.br/aids/pt-br/central-de-conteudo/publicacoes/2026/monitoramento-integrado-do-cuidado-do-hiv-e-da-aids.pdf

20. Latin America and the Caribbean. [cited 20 Aug 2024]. Available: https://www.unaids.org/en/regionscountries/latinamerica

21. Suriname | UNAIDS. [cited 11 Nov 2026]. Available: https://www.unaids.org/en/regionscountries/countries/suriname

22. Stijnberg D, Kee MM, Bergs J, Adhin MR, Schrooten W. The continuum of care for people living with HIV in Suriname: identifying factors influencing the care delivery process. IJQHC Comm. 2022;2:lyac013. doi:10.1093/ijcoms/lyac013

23. Incidence rate of HIV infections | PAHO - PBE website. [cited 11 Nov 2026]. Available: https://pbdigital.paho.org/en/eob-2024-2025/impact-results/16-incidence-rate-hiv-infections

24. Rajasingham R, Medina N, Mousquer GT, Caceres DH, Jordan A, Nacher M, et al. Cost-effectiveness evaluation of routine histoplasmosis screening among people living with advanced HIV disease in Latin America and the Caribbean. PLOS Glob Public Health. 2023;3:e0001861. doi:10.1371/journal.pgph.0001861

25. Duplan H, Rabier S, Sudre C, Adriouch L, Lucarelli A, Huber F, et al. The Evolution of HIV Patient Retention and Care in French Guiana: A Broader View From the Système National des Données de Santé. Front Public Health. 2022;10:823193. doi:10.3389/fpubh.2022.823193

26. Nacher M, Adenis A, Huber F, Hallet E, Abboud P, Mosnier E, et al. Estimation of the duration between HIV seroconversion and HIV diagnosis in different population groups in French Guiana: Strategic information to reduce the proportion of undiagnosed infections. PLoS One. 2018;13:e0199267. doi:10.1371/journal.pone.0199267

